# Cardiometabolic multimorbidity and care experiences in primary healthcare among Brazilian adults aged 50 and over (ELSI-Brazil)

**DOI:** 10.64898/2026.06.16.26355825

**Authors:** Fernando Henrique Aires de Souza, Felipe Mendes Delpino, Sandro Rogério Rodrigues Batista

**Author notes:** Corresponding author: SRRB, Faculty of Medicine, Federal University of Goiás, Goiânia, Brazil.

## Abstract

**Background:** Population aging and the rising burden of non-communicable diseases have increased the prevalence of cardiometabolic multimorbidity (CM-MM) among older adults. Patient-reported experience measures (PREMs) are recognized as essential components of healthcare quality assessment, yet evidence on primary care experiences among individuals with CM-MM remains scarce.

**Objective:** To analyze primary care experiences according to the presence of cardiometabolic multimorbidity among Brazilians aged 50 years and older.

**Methods:** Cross-sectional study using data from the second wave of the Brazilian Longitudinal Study of Aging (ELSI-Brazil, 2019–2021; n = 9,949). CM-MM was defined as the self-reported coexistence of two or more of the following conditions: hypertension, diabetes mellitus, dyslipidemia, acute myocardial infarction, and stroke. Primary care experiences were assessed using a validated 12-item instrument organized into four domains: first-contact access, longitudinality, communication, and care coordination. Associations were estimated using Poisson regression adjusted for sociodemographic, health conditions, and healthcare utilization variables, with stratified analysis by Family Health Strategy (FHS) coverage.

**Results:** **CM-MM** prevalence was 25.5%, with a progressive increase by age and an inverse gradient by education. Individuals with CM-MM reported significantly more positive experiences in longitudinality (mean index 2.53 vs. 2.34; adjusted PR = 1.22; 95%CI 1.12–1.33; p < 0.001) and, to a lesser extent, in communication (mean index 2.68 vs. 2.58; adjusted PR = 1.10; 95%CI 1.00–1.20; p = 0.041). No statistically significant differences were found in first-contact access or care coordination. After stratified by FHS coverage, the observed differences in longitudinality and communication were no longer statistically significant.

**Conclusions:** **CM-MM** was associated with more positive primary care experiences in longitudinality and communication. The absence of differentiated experiences in first-contact access and coordination highlights structural gaps in primary care responsiveness to individuals with greater clinical complexity.

## BACKGROUND

Population ageing is occurring against a backdrop of profound social, regional and healthcare-access inequalities, posing increasing challenges for health systems and, in particular, for primary care (PC) [1,2,3,4,5]. In this context, the occurrence of multiple simultaneous chronic conditions in the same individual, defined as multimorbidity, has become increasingly prevalent in health services, adding substantial complexity to clinical management [3,4,6]. Among the different multimorbidity patterns, cardiometabolic multimorbidity (CM-MM) occupies a central position, grouping conditions such as hypertension, diabetes mellitus, cardiovascular disease, stroke, dyslipidaemia and obesity — all highly prevalent conditions [7,8,9,10,11,12,13] that frequently co-occur and are associated with increased risks of disability [4,14], hospital admission [15,16], intensive use of health services [16,17] and mortality [7,10,11,18].

Primary care plays a strategic role in the care of people with multiple chronic conditions, serving as the preferred point of entry to the health system and being responsible for longitudinal care, care coordination, prevention of complications and links with other levels of care [2,5,19,20,21]. For people living with CM-MM, primary care experiences may involve frequent consultations [16,17], continuous use of medications — often including polypharmacy [22,23] — more intensive clinical monitoring and diagnostic testing [21], self-care support [23] and referrals to other levels of care [21]. In this regard, patient-reported experiences in primary care services can reveal the extent to which care responds to the complex needs associated with CM-MM, including first-contact access, longitudinality (continuity), communication and care coordination [24,25,26].

Although primary care plays a central role in well-performing health systems, individuals with multimorbidity, including those with CM-MM, may face substantial difficulties in their care [3]. Service organisation is often still guided by single-disease approaches, fragmented protocols and episodic responses, which may be insufficient to meet the complex needs of individuals with CM-MM [5,21,27]. Furthermore, social inequities may shape distinct care experiences [3,17], particularly among older adults [3], a group marked by considerable heterogeneity in living conditions [3] and in coverage by health services [19].

Although the literature has advanced in describing the prevalence, patterns and outcomes associated with CM-MM, studies examining how its specific occurrence relates to patient experiences in primary care services remain scarce. Most investigations focus on disease counts, service utilisation or clinical outcomes, while dimensions such as access, longitudinality, communication and care coordination are less frequently explored [17,19,26,21]. This gap is relevant because patient experience is a fundamental dimension of care quality and can reveal organisational weaknesses that are not captured by clinical or administrative indicators alone [24,25].

In this context, this study aims to analyse primary care experiences according to the presence of CM-MM among Brazilians aged 50 years and over, using data from the Brazilian Longitudinal Study of Ageing (ELSI-Brazil). By investigating this association in a nationally representative sample, the study seeks to contribute to understanding the care needs of people with CM-MM and to strengthening more integrated, longitudinal, equitable and person-centred primary care, capable of responding to the challenges imposed by population ageing and the growing burden of non-communicable diseases in Brazil.

## METHODS

This study followed the STROBE (Strengthening the Reporting of Observational Studies in Epidemiology) checklist for cross-sectional studies to ensure transparent and rigorous reporting [28].

### Setting and data

ELSI-Brazil is a nationally representative, household-based longitudinal study designed to monitor the health, functional and socioeconomic conditions of adults aged 50 years and over living in the Brazilian community. It is the largest longitudinal research initiative on ageing conducted in Brazil and forms part of the international network of population ageing studies, such as the Health and Retirement Study (HRS) in the United States and the Survey of Health, Ageing and Retirement in Europe (SHARE). The first wave of ELSI-Brazil was conducted between 2015 and 2016, and the second wave, used in the present study, between 2019 and 2021. The study is coordinated by the René Rachou Research Centre of the Oswaldo Cruz Foundation (Fiocruz Minas) and provides data representative of the Brazilian population aged 50 years and over living in urban and rural areas across all regions of the country.

### Variables

#### Primary care experiences

Participants’experiences of PC were measured using a composite index of 12 self-reported indicators, grouped into four core domains: first-contact access, longitudinality, communication, and coordination {12]. Each item reflects an experience (positive or negative) with the health care provider, such as ease of making appointments, continuity with the same doctor, careful listening, clear explanations, and support in coordinating specialist care.

Responses were given on a Likert scale and dichotomized, classifying “always” or “most of the time” as positive and “rarely” or “never” as negative. The total score, ranging from 0 to 12, represents the number of positive primary care attributes perceived—where higher values indicate better PHC experience. The index was calculated only for participants who answered at least two items per domain, ensuring response consistency.

A composite Primary Care Effectiveness Index was calculated as the sum of positive primary care attributes perceived by the participant across all 12 indicators, generating a continuous scale ranging from 0 to 12. Each domain contributes a sub-index ranging from 0 to 3, corresponding to the number of positive attributes reported within that domain.

### Cardiometabolic multimorbidity (CM-MM)

The presence of CM-MM was evaluated using a list of 16 physical and mental morbidities, as follows: 1) hypertension, 2) diabetes, 3) high cholesterol, 4) myocardial infarction 5) stroke. All conditions were evaluated using a self-reported medical diagnosis. The question “*Has a physician already diagnosed you as having (each disease)?*” was asked in relation to each disease.

### Covariables

We included the following covariates in the analyses: sex (male; female); ge (by group: 50–59; 60–69; 70–79; 80 years and over); education level (1–4; 5–8; 9–11; more than 12 years of study); marital status (married; not married); lives alone (yes; no); area of residence (urban; rural); Brazilian geopolitical region (North; Northeast; Southeast; South; Centre-West); have private health insurance (yes; no); have coverage by the Family Health Strategy (yes; no). The following covariates related to health conditions of importance in the older population were considered: self-rated health (very good/good; fair; poor/very poor); polypharmacy (five or more medications) (yes; no); falls in the past 12 months (yes; no); hospitalisation in the past 12 months (yes; no).

### Statistical analysis

All analyses took into account the complex sampling design of ELSI-Brazil, incorporating sampling weights, strata and primary sampling units to generate estimates representative of the target population. Categorical variables were described as weighted frequencies and proportions, and continuous primary care domain indices as weighted means and standard deviations.

Cardiometabolic multimorbidity was operationalised in:

● CM-MM: ≥2 of the following conditions — hypertension, diabetes, high cholesterol, myocardial infarction and stroke.

The prevalence of CM-MM and its association with sociodemographic and health-related characteristics were estimated using Poisson regression with robust variance, an approach recommended for common binary outcomes. Results are presented as prevalence ratios (PRs) with 95% confidence intervals (95%CI).

To assess the association between CM-MM and primary care experiences, Poisson regression models with robust variance were fitted for each of the four domain indices — first-contact access, longitudinality, communication and care coordination — and for each individual indicator, in crude and adjusted models. Adjusted models included sex, age group, education, marital status, living alone, area of residence, geopolitical region, private health insurance and FHS coverage. Covariables were selected a priori based on their theoretical relevance as potential confounders, informed by the literature. Results are expressed as adjusted prevalence ratios (aPRs) with 95%CI, and associations were considered statistically significant when the 95%CI did not include 1.0.

To explore whether this association differed according to the care model, the analytic sample was restricted to individuals with CM-MM and we examined whether primary care experiences varied according to FHS coverage. For this purpose, Poisson models with robust variance were fitted including the same covariables as in the main analyses, except FHS coverage itself, which was the exposure of interest.

Statistical significance was defined as p < 0.05. All analyses were conducted using Stata SE 15.0 (StataCorp, College Station, TX, USA).

## RESULTS

The final sample comprised 9,949 individuals aged 50 years and over, of whom 2,537 (25.5%) presented cardiometabolic multimorbidity. The distribution of sociodemographic and health characteristics according to the presence or absence of CM-MM is shown in Table 1. CM-MM was more prevalent among women (55.1% of cases), displayed a progressive and statistically significant increase with advancing age — from 37.2% in the 50–59 age group to 9.8% among those aged 80 years or over — and showed an inverse association with education, with progressively lower prevalence ratios in the highest schooling categories (PR = 0.68; 95%CI 0.52–0.89 for ≥12 years of study). Living alone (PR = 1.24; 95%CI 1.02–1.50), poorer self-rated health (PR = 2.37; 95%CI 1.99–2.82 for poor/very poor health), polypharmacy (PR = 1.78; 95%CI 1.54–2.04) and hospitalisation in the previous 12 months (PR = 1.50; 95%CI 1.25–1.80) were also significantly associated with CM-MM. FHS coverage and possession of private health insurance did not show statistically significant associations with the occurrence of CM-MM.

**Table 1.**
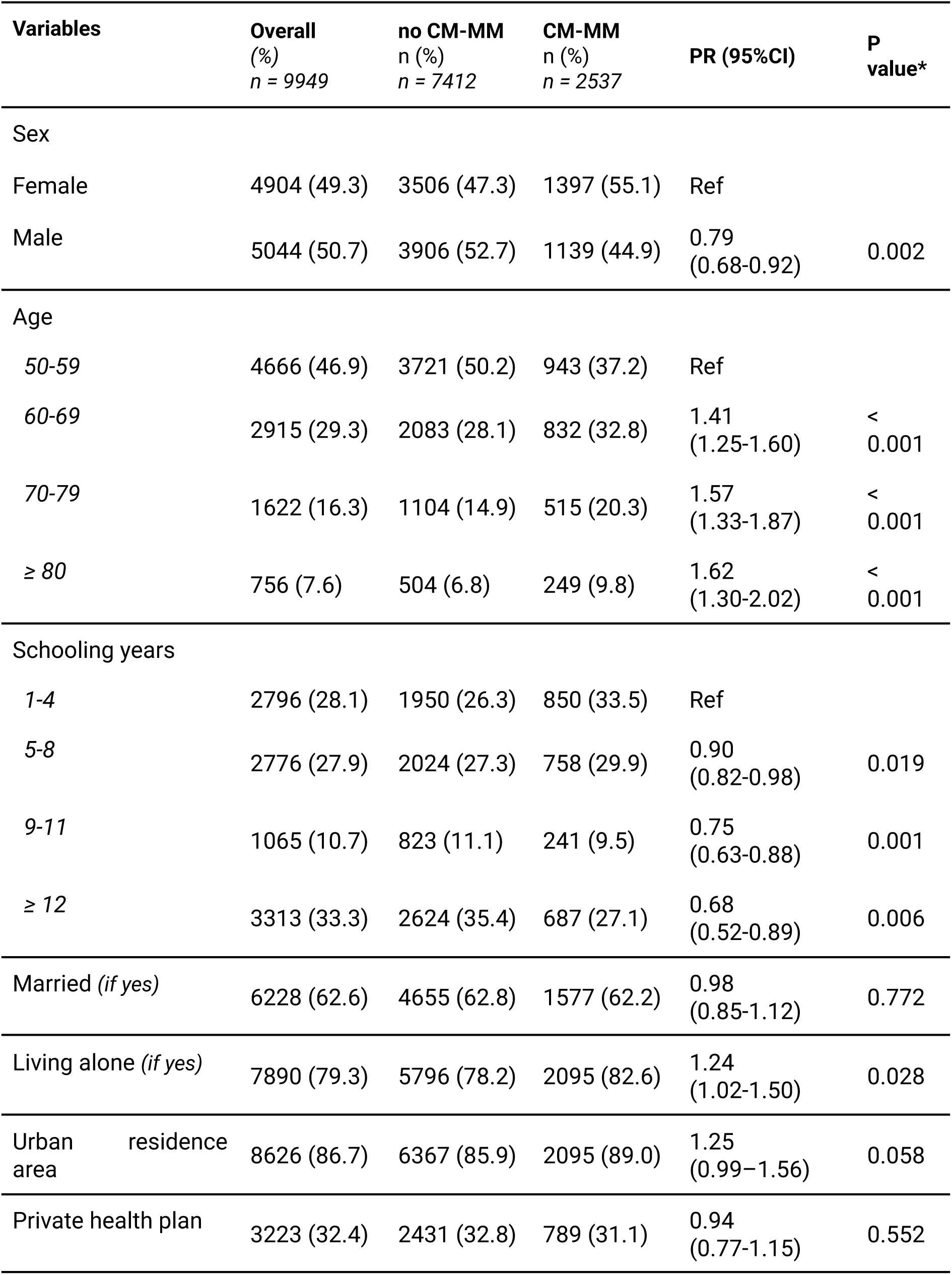
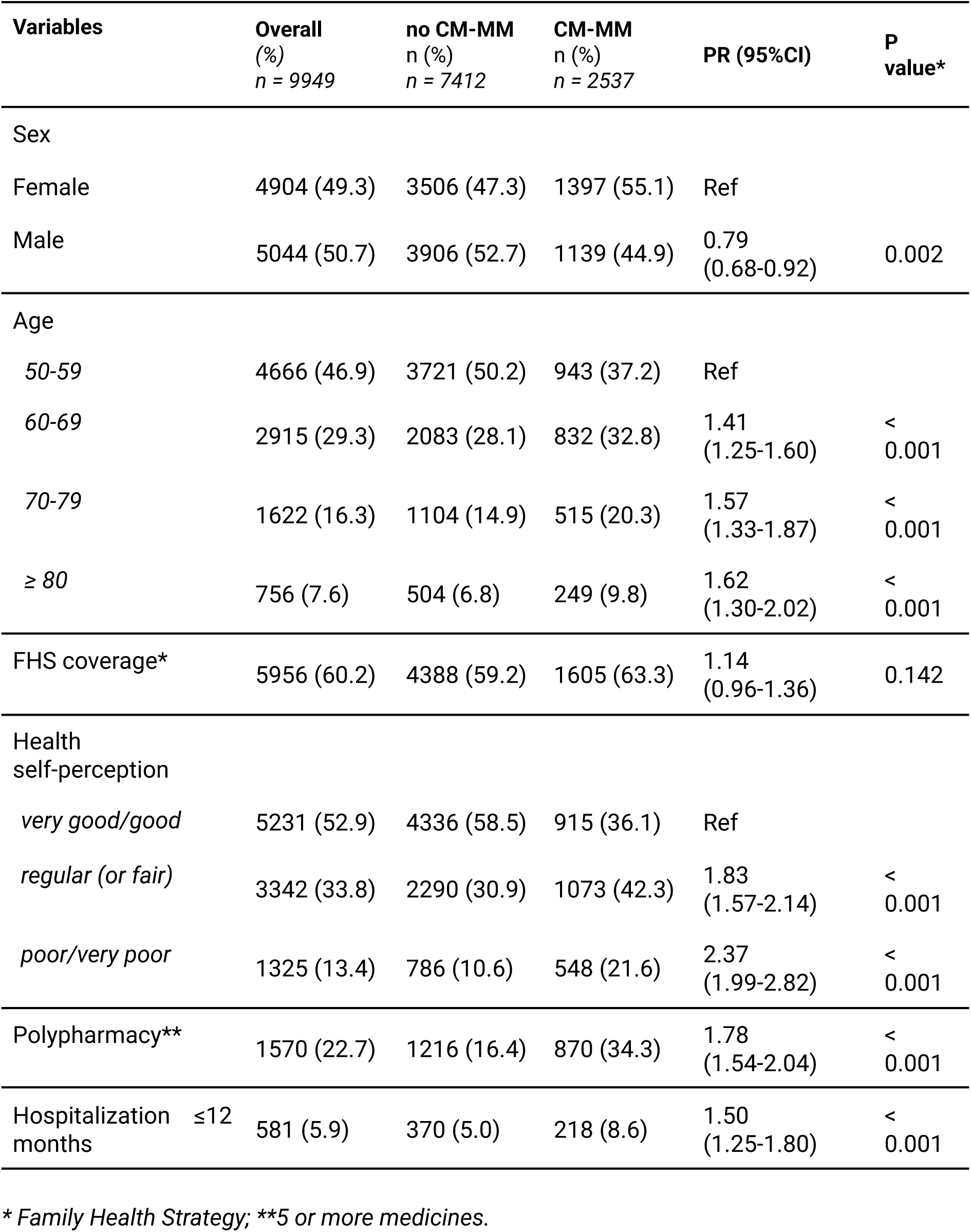
Distribution of the overall sample and according to the presence of cardiometabolic multimorbidity (CM-MM), ELSI-Brazil, second wave, 2019–2020.

Table 2 presents the distribution of the investigated morbidities according to the presence of CM-MM. Among the conditions composing the construct adopted in this article, systemic arterial hypertension showed the strongest association (PR = 12.15; 95%CI 9.63–15.33), followed by high cholesterol (PR = 6.67; 95%CI 5.74–7.74), diabetes mellitus (PR = 5.20; 95%CI 4.51–6.01), acute myocardial infarction (PR = 3.65; 95%CI 3.17–4.22) and stroke (PR = 3.50; 95%CI 3.02–4.05). Among morbidities not included in the CM-MM construct, significant associations were observed for heart failure, angina, arthritis or rheumatism, osteoporosis, back problems, ophthalmological problems, depression, asthma and chronic kidney failure. Cancer, chronic pulmonary conditions, Parkinson’s disease and Alzheimer’s disease did not present statistically significant associations with CM-MM.

**Table 2.** Morbidities in overall sample and according to the presence of cardiometabolic multimorbidity (CM-MM), ELSI-Brazil, second wave, 2019–2020.

| Variables | Overall<br>(%)<br>*n* = 9949 | no CM-MM<br>*n* (%)<br>*n* = 7412 | CM-MM<br>*n* (%)<br>*n* = 2537 | PR (95%CI) | P<br>value* |
| --- | --- | --- | --- | --- | --- |
| Hypertension | 4676<br>(47.0) | 2372<br>(32.0) | 2321<br>(91.5) | 12.15<br>(9.63-15.33) | <<br>0.001 |
| Diabetes | 1652<br>(16.6) | 385 (5.2) | 1289<br>(50.8) | 5.20 (4.51-6.01) | <<br>0.001 |
| High cholesterol | 2189<br>(22.0) | 608 (8.2) | 1657<br>(65.3) | 6.67 (5.74-7.74) | <<br>0.001 |
| Heart attack | 358 (3.6) | 59 (0.8) | 304 (12.0) | 3.65 (3.17-4.22) | <<br>0.001 |
| Heart failure | 418 (4.2) | 193 (2.6) | 233 (9.2) | 2.29 (1.87-2.79) | <<br>0.001 |
| Angina | 179 (1.8) | 82 (1.1) | 101 (4.0) | 2.23 (1.60-3.10) | <<br>0.001 |
| Stroke | 358 (3.6) | 67 (0.9) | 292 (11.5) | 3.50 (3.02-4.05) | <<br>0.001 |
| Asthma | 298 (3.0) | 185 (2.5) | 112 (4.4) | 1.49 (1.14-1.94) | 0.004 |
| Pulmonary problems* | 358 (3.6) | 245 (3.3) | 109 (4.3) | 1.22 (0.96-1.55) | 0.097 |
| Arthritis<br>or<br>rheumatism | 1671<br>(16.8) | 1045<br>(14.1) | 632 (24.9) | 1.64 (1.42-1.88) | <<br>0.001 |
| Osteoporosis | 1015<br>(10.2) | 630 (8.5) | 383 (15.1) | 1.57 (1.34-1.85) | <<br>0.001 |
| Back problems | 2995<br>(30.1) | 2061<br>(27.8) | 939 (37.0) | 1.36 (1.19-1.56) | <<br>0.001 |
| Depression | 1214<br>(12.2) | 823 (11.1) | 396 (15.6) | 1.32 (1.09-1.61) | 0.005 |
| Cancer | 537 (5.4) | 400 (5.4) | 140 (5.5) | 1.01 (0.74-1.39) | 0.936 |
| Chronic renal failure | 229 (2.3) | 141 (1.9) | 89 (3.5) | 1.51 (1.13-2.01) | 0.005 |
| Parkinson's disease | 90 (0.9) | 59 (0.8) | 28 (1.1) | 1.17 (0.70-1.96) | 0.541 |
| Alzheimer's disease | 109 (1.1) | 74 (1.0) | 36 (1.4) | 1.22 (0.79-1.90) | 0.370 |
| Ophthalmological problems** | 3273 (32.9) | 2224 (30.0) | 1050 (41.4) | 1.44 (1.25-1.66) | < 0.001 |

\* *Emphysema, chronic bronchitis or chronic obstructive pulmonary disease - COPD*
\*\* *Cataract, glaucoma, diabetic retinopathy, macular degeneration.*

Figure 1 shows the distribution of positive primary care experiences according to the presence of CM-MM. Individuals with CM-MM reported significantly more positive experiences in the domains of longitudinality and communication compared with those without CM-MM, whereas no statistically significant differences were identified in the domains of first-contact access and care coordination. The Primary Care Effectiveness Index by CM-MM status is presented in Figure 3, which shows that the largest differences between groups were observed in the longitudinality domain.

**Figure 1.**
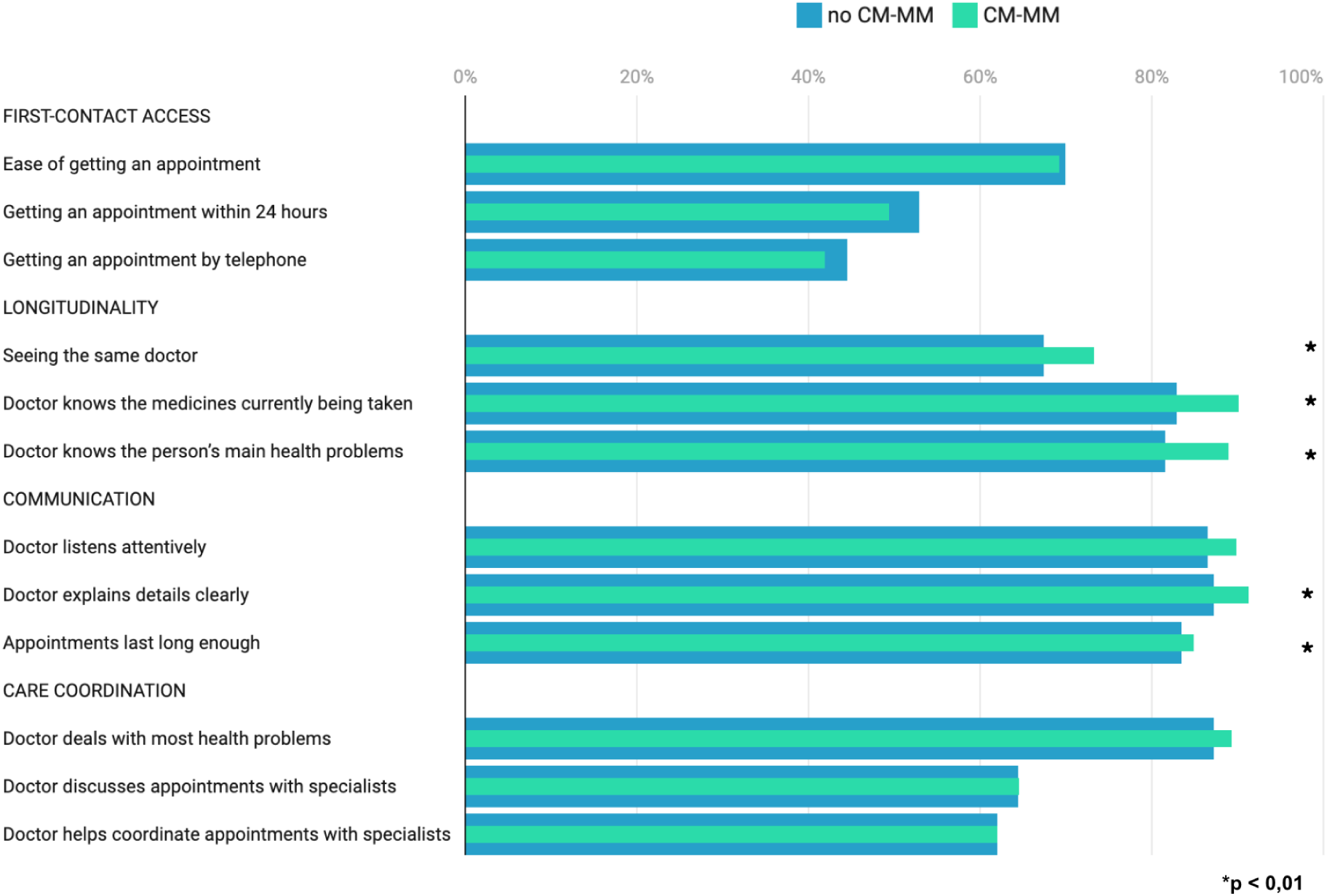
Positive experiences in primary care according to the presence of cardiometabolic multimorbidity (CM-MM), ELSI-Brazil, second wave, 2019–2021.

**Figure 2.**
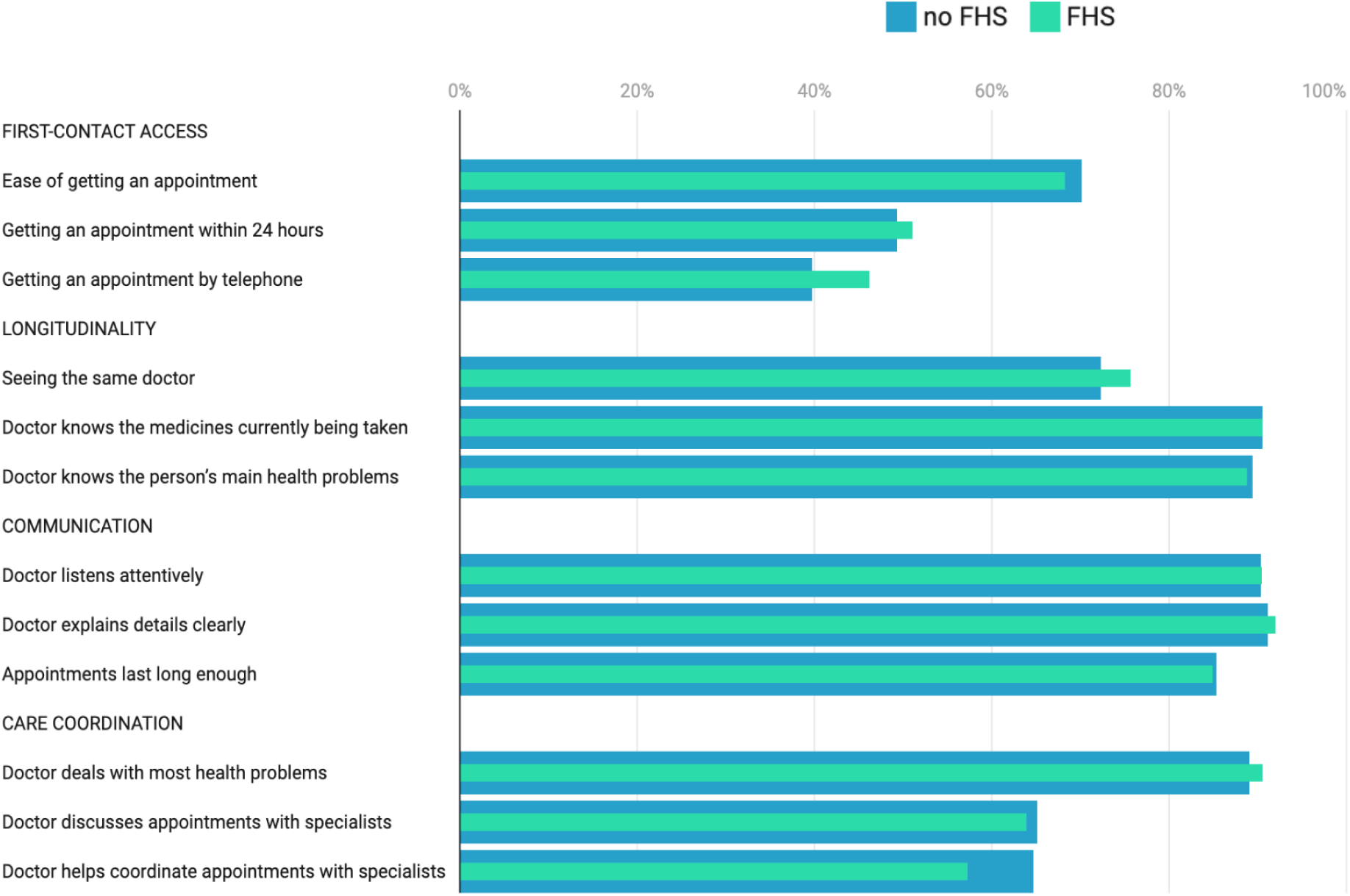
Positive experiences in primary care among individuals aged 50 years and over with cardiometabolic multimorbidity (CM-MM), according to FHS coverage, ELSI-Brazil, second wave, 2019–2021.

**Figure 3.**
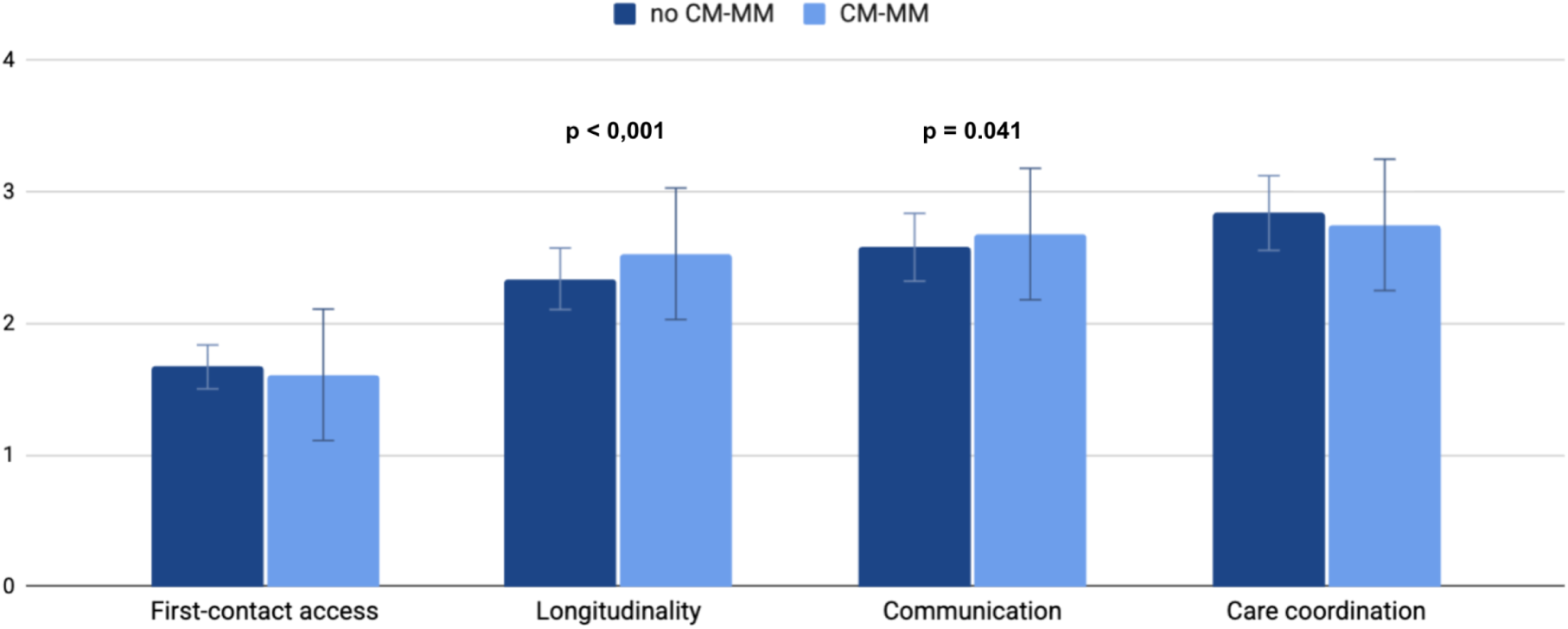
Primary care effectiveness index according to the presence of cardiometabolic multimorbidity (CM-MM), ELSI-Brazil, second wave, 2019–2021.

Figure 2 presents positive primary care experiences among individuals with CM-MM according to FHS coverage. In the stratified analysis, the differences previously observed in the domains of longitudinality and communication between those with and without CM-MM were no longer statistically significant, suggesting a potential equalising effect of the FHS care model. Figure 4 shows the Primary Care Effectiveness Index according to FHS coverage among individuals with CM-MM, demonstrating no statistically significant differences between the two groups across any of the evaluated domains.

**Figure 4.**
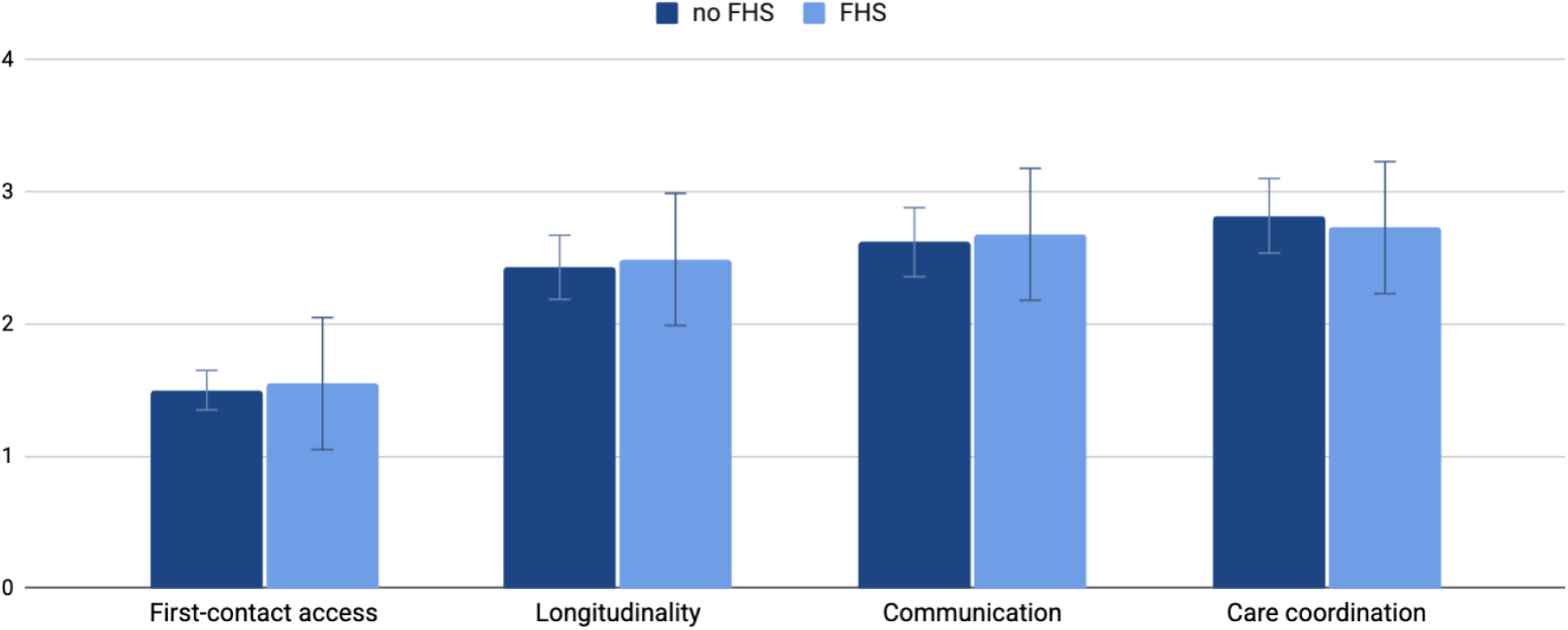
Primary care effectiveness index according to the Family Health Strategy (FHS) coverage among individuals with cardiometabolic multimorbidity (CM-MM), ELSI-Brazil, second wave, 2019–2021.

## Discussion

The epidemiological findings of this study are consistent with patterns described in the national and international literature on CM-MM. The prevalence of 25.5%, with a progressive gradient across age groups and an inverse association with education, reproduces the pattern observed in population-based studies [5,6,15]. The higher prevalence among women (55.1%) contrasts with North American and Brazilian studies that report a predominance among men [6,15], a difference that may reflect the inclusion of hypertension and dyslipidaemia in the construct adopted here — conditions with higher self-reported prevalence among Brazilian women. The strong associations with polypharmacy (PR = 1.78), negative self-rated health (PR = 2.37) and hospitalisation (PR = 1.50) reflect the clinical and functional burden imposed by the coexistence of multiple cardiometabolic conditions, in line with evidence documenting the multiplicative impact of CM-MM on mortality, disability and intensive service use [5,7,9].

The broad morbidity profile associated with CM-MM — with significant associations with depression, arthritis, osteoporosis, chronic kidney disease and ophthalmological problems — reinforces the systemic nature of this condition and the challenges it poses to care models that remain organised around single diseases [3,14,22]. Cardiometabolic conditions rarely occur in isolation; they frequently coexist with discordant conditions across multiple organ systems, amplifying clinical complexity and treatment burden [31].

In the domain of first-contact access, the absence of differences between groups — with a Primary Care Effectiveness Index of 1.61 (on a 0–3 scale) for people with CM-MM — indicates that access difficulties are transversal and affect all primary care users indiscriminately, regardless of morbidity profile. This result is consistent with literature describing access as an attribute conditioned by organisational and structural features of services — opening hours, appointment availability, demand management — rather than by users’ clinical characteristics [16,17]. The lack of differentiation in system response suggests that individuals with greater clinical complexity face the same structural barriers as other users, which may mask more severe unmet needs.

The longitudinality domain concentrated the most robust associations in this study, with all three indicators showing statistically significant differences after adjustment and the Primary Care Effectiveness Index consistently higher among people with CM-MM (2.53 versus 2.34; aPR = 1.22; 95%CI 1.12–1.33; p < 0.001). This finding is coherent with the hypothesis that the chronic nature of cardiometabolic conditions intensifies longitudinal contact with primary care services — prescription renewals, monitoring of clinical parameters, therapeutic adjustments — creating conditions for the development of relational bonds over time [12,16]. The indicator with the strongest association was the doctor’s knowledge of the medications in use (aPR = 1.69; 95%CI 1.35–2.11), a finding of particular clinical relevance in a group with high prevalence of polypharmacy. These results are supported by evidence that relational continuity is the attribute most valued and actively sought by patients with complex chronic conditions, precisely because they perceive that its absence compromises the quality and safety of care [34].

In the communication domain, the Primary Care Effectiveness Index was also higher among individuals with CM-MM (2.68 versus 2.58; aPR = 1.10; 95%CI 1.00–1.20; p = 0.041), driven mainly by the indicator capturing the clarity of the doctor’s explanations (aPR = 1.38; 95%CI 1.10–1.72; p = 0.005). The asymmetry between indicators — with attentive listening losing statistical significance after adjustment and consultation length showing no difference in any analysis — is consistent with findings that describe explanatory communication as more systematically practised in encounters with patients of greater clinical complexity, while high-quality listening remains more dependent on individual relational competences of the professional [35,36].

The analysis stratified by FHS coverage showed that the differences observed in longitudinality and communication disappeared completely within the Strategy, with Primary Care Effectiveness Index values almost identical between those with and without CM-MM, both among individuals covered and not covered by the FHS. This equalising effect may be explained by two complementary mechanisms. The first is structural: the FHS actively institutionalises longitudinality through population empanelment, the work of community health workers and the establishment of multiprofessional reference teams [12,26]. The second is methodological: Primary Care Effectiveness Index scores within the FHS approach the maximum value for each domain — a ceiling effect that compresses score distributions and limits the instrument’s ability to detect differences between subgroups, a phenomenon recognised in the PREMs literature [23]. The multiprofessional nature of the FHS is aligned with conditions identified in the international literature as favourable for team-based work to strengthen, rather than fragment, relational continuity [26].

In the care-coordination domain, the absence of differences between groups — with high Primary Care Effectiveness Index scores in both (2.75 and 2.84) — represents a paradoxical finding. People with CM-MM have objectively greater needs for coordination across different points of the care network, yet they report experiences indistinguishable from those without multimorbidity. Three mechanisms may explain this paradox. The first is an adapted-expectations bias: populations historically exposed to fragmented systems may have developed expectations calibrated to insufficiency, rating as positive a perceived level of coordination that, in more integrated systems, would be considered unsatisfactory [17,23]. The second is a respondent-selection effect: individuals whose conditions are managed entirely within primary care, without the need for specialist referral, tend to rate coordination positively regardless of how well counter-referral mechanisms function, thereby diluting the negative signal from those who have effectively experienced fragmentation. The third is a conceptual limitation of the instrument: coordination items capture the doctor’s coordinating intent at the point of contact, rather than the systemic functioning of integration between levels of care — a dimension that would require assessment of system structures and processes, not solely users’ self-reported experience [17].

This study has limitations that should be taken into account when interpreting the findings. Data collection for the second wave of ELSI-Brazil (2019–2021) was interrupted by the SARS-CoV-2 pandemic, which may have influenced participants’ perceptions of health services. The cross-sectional design precludes establishing causal relationships between CM-MM and care experience. The instrument has a physician-centred focus, underestimating continuity provided by other members of the multiprofessional team — a particularly relevant limitation in the FHS context, where nurses and community health workers often constitute the first longitudinal point of contact [26]. The instrument also does not capture comprehensiveness as a dimension of care experience [17]. Additionally, self-reported data are subject to recall, acquiescence and survival biases, which are well recognised in the PREMs literature [23–25].

### Conclusions

This study showed that cardiometabolic multimorbidity is associated with significantly more positive experiences in the domains of longitudinality and, to a lesser extent, communication in primary care among Brazilian adults aged 50 years and over. The absence of differences in first-contact access and care coordination indicates that the system does not differentiate its response according to the greater clinical complexity of this group, revealing gaps that warrant attention in policies to strengthen primary care.

The equalising effect observed in the stratified analysis by the Family Health Strategy — in which individuals with and without CM-MM exhibited similarly high longitudinality scores on the Primary Care Effectiveness Index — raises the question of whether the model responds proportionally to the greater clinical complexity of this group, or instead treats individuals with objectively different needs in the same way. Determining whether the FHS overcomes or reproduces this pattern requires future studies capable of assessing not only perceived experience, but also the adequacy and proportionality of the care response to clinical complexity.

These findings reinforce the importance of systematically incorporating instruments to measure care experience — validated PREMs applied in population-based surveys — as a permanent component of primary care quality assessment in Brazil, recognising that the user perspective captures dimensions of care quality that traditional clinical and administrative indicators do not. Longitudinal studies using future ELSI-Brazil waves, instruments able to assess continuity with the multiprofessional team as a whole, and investigations of the therapeutic pathways of people with CM-MM in the Brazilian health system constitute priority research agendas for advancing understanding of the needs of this growing population group.

## Data Availability

Availability of data and materials: All ELSI-Brazil Study data are available from the study website <https://elsi.cpqrr.fiocruz.br/data-access/>;

https://elsi.cpqrr.fiocruz.br/data-access

## Declarations

### Ethics approval and consent to participate

ELSI-Brazil was approved by the Fundação Oswaldo Cruz (FIOCRUZ) ethics committee, Minas Gerais, Brazil (protocol number 34649814.3.0000.5091).

### Consent for publication

Not applicable.

### Availability of data and materials

All ELSI-Brazil Study data are available from the study website <**Error! Hyperlink reference not valid.**>

### Authors’ Contributions

Research designed by FHAS and SRRB (project development, objectives, research methods). FMD and SRRB conducted data analysis. FHAS and SRRB wrote the article and were mainly responsible for the final content. All authors assisted in the interpretation of results and critical revision of the manuscript. All authors contributed to the final manuscript and approved the final version.

## Acknowledgements

We also thank the individuals who participated in the ELSI-Brazil study.

